# Using MRI whole brain atrophy and clinically reported outcomes in combination to assess interim treatment response in multi-arm multi-stage trials in progressive multiple sclerosis

**DOI:** 10.64898/2026.06.26.26356667

**Authors:** Matthew Burnell, Jennifer Nicholas, Rachel Burton, Sean Apap Mangion, Jeremy Chataway, James Carpenter

**Author notes:** **Correspondence** Corresponding author: Matthew Burnell, UCL Innovative Clinical Trials Unit, 90 High Holborn, London, UK.

## Abstract

**Background:** Interim stage outcomes in multi-arm multi-stage trials need not be the same as the final primary outcome, and should be selected with the goal of providing the best chance of continuing with an effective treatment in the study during the early stage where there is also potential to drop an ineffective arm. Jointly considering multiple outcomes can enhance that ability to detect an emerging signal.

**Methods:** The Optimal Clinical Trials Platform for Progressive Multiple Sclerosis (OCTOPUS) study is a randomised, placebo-controlled, double-blind, phase 3, MAMS trial testing treatments for people with progressive multiple sclerosis. The interim analysis was to be based solely on an MRI outcome; reduction in whole brain atrophy rate. Accumulating data from external trials led to concern that this outcome may result in prematurely rejecting an effective treatment. As a solution we propose adding 3 clinical outcomes to the MRI outcome and propose a multivariate mixed model to accommodate them jointly, despite significant differences in scale and even measurement type. We show how use of the model-derived covariances allows us to linearly combine the treatment effects of disparate outcomes into a single treatment effect. We also describe how to analytically calculate power for this combination and compared it to the individual components’ performance.

**Results:** Based on variance data from a previous study, we found power was increased moderately by 7%, from 83% to our target 90% given the assumed respective treatment effect sizes for OCTOPUS. When considering observed effect sizes from other trials these power improvements were maintained, despite there being great variability between the outcome effects.

**Conclusions:** Whilst not greatly boosting power, we argue that this strategy improves the interim outcome measure by also making it more resilient to the uncertainty surrounding effect size, and mitigating against unexpected negative results by spreading the liability across related but distinct outcomes.

## 1 INTRODUCTION

The advantages of multi-arm multi-stage (MAMS) designs, initially demonstrated in the successful Phase 3 prostate cancer trial STAMPEDE ^1^, have led to their application across a range of diseases. To support their use in neurodegenerative diseases, in 2021 the Medical Research Council (MRC) Clinical Trials Unit (CTU), now known as UCL Innovative CTU (InCTU), established A Collaboration of groups Running and reporting MAMS trials in neurodegenerative Diseases (ACORD). ACORD is now supporting ongoing MAMS trials in motor neuron disease ^2^, progressive multiple sclerosis (PMS) ^3,4^, Parkinson’s disease ^5^, and Alzheimer’s Disease (AD SMART).

A key component of MAMS designs is the option to stop intervention arms early if there is insufficient evidence of activity against the disease. This decision is typically not based on the primary outcome measure in the trial, nor necessarily a formal surrogate endpoint, but chosen from a range of early measures (e.g. tumour progression in cancer), across which we would expect to see early signs of activity if the drug is ultimately going to show clinical benefit.

Figure 1 shows a the design schematic for OCTOPUS. In the early stages of a MAMS trial, a typically non-binding statistical test is used to guide the decision to continue a candidate therapy into the next stage. With the selected test having high power (say ≥ 90%), but also high probability of a type 1 error (typically ≥ 10%). These values are chosen to minimize the probability of discontinuing interventions that may subsequently prove clinically effective. As the trial progresses, the subsequent stages retain a high power, while the probability of a type-1 error is lowered. The hurdle for interventions to clear in order to continue is therefore gradually raised until the final stage, in which the primary trial outcome is assessed using typical error rates.

**FIGURE 1.**
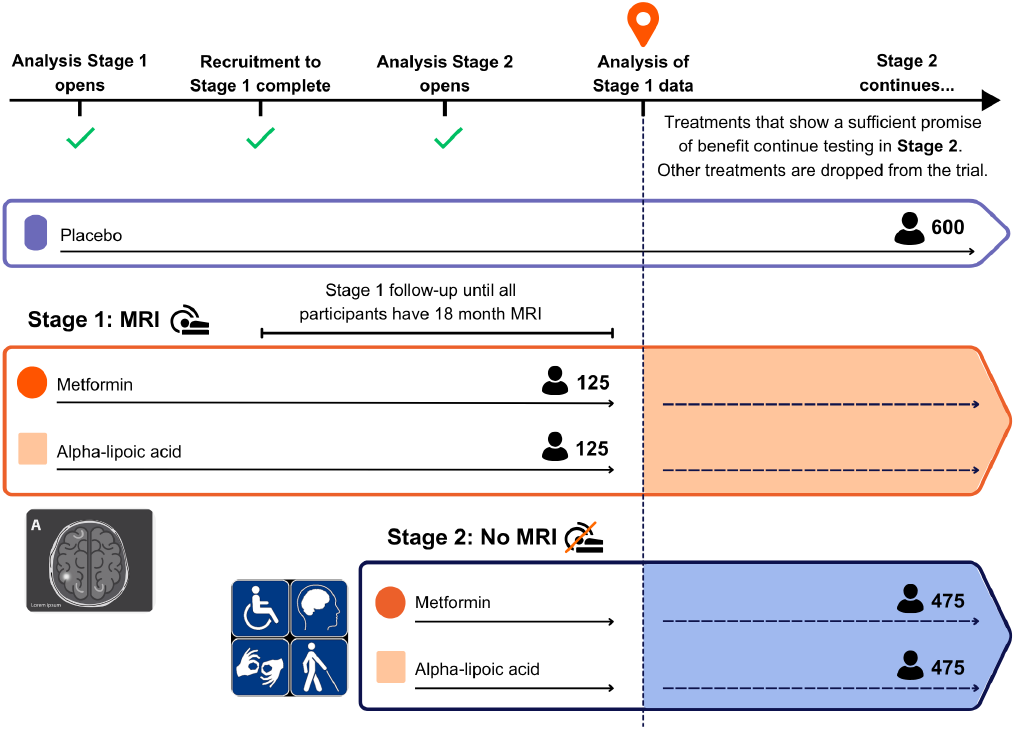
Schematic representation of the OCTOPUS MAMS trial in progressive multiple sclerosis comparing two experimental arms to standard of care in two stages.

To date, most MAMS trials have used a single outcome measure, at least per analysis stage. In this paper we describe a general approach showing how multiple (continuous) outcomes may be considered jointly for the purposes of increasing power and defraying the risk of using a single a-priori choice of outcome. The remainder of the article is structured as follows. In Section 2 we describe the MAMS study that motivated this work. In Section 3, we outline our proposed statistical methods for incorporating information from multiple outcome measures in early stage analyses. In Section 4, we present the results when these methods are applied to the design of a two-stage MAMS study and in Section 4.2 examine how they would have performed if applied to previous phase 3 trials in PMS. Finally, in Section 5, we provide a discussion and compare our findings with how other trials have been designed.

## 2 MOTIVATION: OCTOPUS TRIAL

The Optimal Clinical Trials Platform for Progressive Multiple Sclerosis (OCTOPUS) study is an international randomised, placebo-controlled, double-blind, phase 3, MAMS trial testing treatments for people with PMS. The key eligibility criteria are age 25 to 70 years (inclusive) with either primary or secondary PMS with an Expanded Disability Status Score (EDSS) score of 4.0 to 8.0 (inclusive). Steady progression must be the major cause of increasing disability rather than relapse in the preceding 2 years. Participants are randomly allocated in 1:1:1 ratio between three arms: racemic alpha lipoic acid (R/S ALA); immediate release metformin; and placebo, with all participants on best standard of care, including disease modifying treatment if appropriate.

The OCTOPUS trial will evaluate drug efficacy by comparing each of the active arms to placebo in a two-stage design. Analysis Stage 1 will assess efficacy (i.e. signs of drug activity against the disease), while Analysis Stage 2 will be the final primary outcome analysis. The OCTOPUS primary outcome, which will be evaluated at the end of Stage 2, is 6-month confirmed disability progression (CDP) in the composite EDSS-Plus, consisting of CDP on one or more of three clinically reported outcomes (CROMS): (i) the EDSS, (ii) Timed 25-Foot Walk test (T25FW), and (iii) 9-Hole Peg Test (9HPT). ^6^

The rate of whole brain atrophy (WBA), measured from Magnetic Resonance Imaging (MRI), was selected as the Analysis Stage 1 interim outcome in the original design of OCTOPUS ^3,4^. This outcome was chosen because it was established as a primary outcome in phase 2 trials in PMS ^7–9^, reflects underlying neuroaxonal loss associated with future clinical disability ^10,11^, and is associated with treatment effects seen in clinical outcomes in previous trials. In the original OCTOPUS trial design, it was calculated that after allowing for drop-out, 125 patients per arm with MRI would provide 95% power to detect a 0.15%/year reduction in the rate of WBA at 18 months, with a one-sided alpha of 0.35, under the assumption that the standard deviation of the atrophy rate will be 0.55%/year. This choice of power and type-1 error are typical for early stage analyses in MAMS studies, when we wish to continue if there is any indication of drug activity against the disease.

Despite the strong rationale for use of WBA as an interim outcome in adaptive trials in PMS, accumulating data suggests it may not be sufficient to use WBA alone ^12,13^. Phase 3 trials demonstrating a benefit of treatment in slowing disability progression in PMS have not always shown concordant results on WBA. For example, both the ORATORIO trial ^14^ and later ORATORIO-HAND ^15^ trial demonstrated a reduction in disability progression with ocrelizumab compared to placebo (hazard ratio (HR) for CDP: 0.76 and 0.70 respectively). However, the 17.5% relative reduction in WBA rates in ORATORIO was not replicated in ORATORIO-HAND, which found minimal differences in atrophy rates between ocrelizumab and placebo (mean difference -0.002%/year) (Table 1). Similarly, the HERCULES ^16^ trial demonstrated a positive treatment effect for tolebrutinib (HR for CDP of 0.69) with a non-significant difference in WBA ^13^. Discrepancies like this between an interim WBA outcome and a CDP primary outcome increase the probability of discontinuing interventions that may subsequently prove clinically effective. For example, if an effective treatment reduced the WBA rate by 0.05%/year instead of the originally assumed 0.15%/year then power at Analysis Stage 1 in OCTOPUS would drop from 95% to 63%. Therefore, there is a clear need to supplement whole brain atrophy with further outcome data for use in interim decision-making in MAMS trials in PMS.

**TABLE 1.**
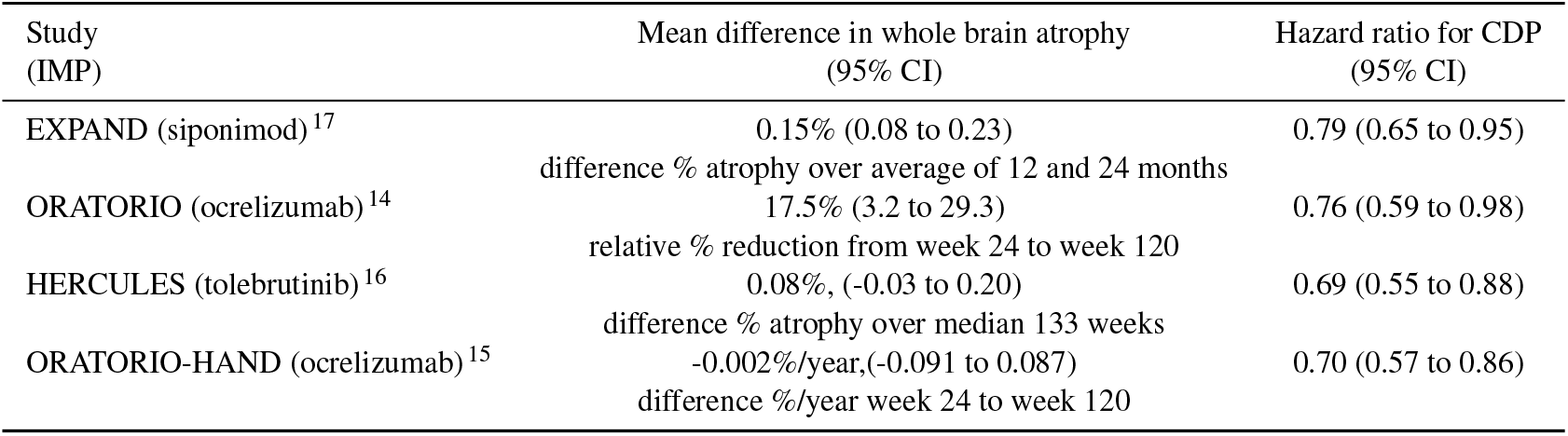
Summary of findings on CPD and whole brain atrophy from four positive phase 3 trials in PMS.

## 3 METHODS

We propose a method for combining data from multiple outcomes for interim assessment in MAMS trials. Specifically, we describe a joint multivariate mixed effects linear regression model for the three CROMS measures and brain atrophy. As we will see, this requires a non-standard random effects covariance structure.

### 3.1 Dataset

We demonstrate our approach using data from the MS-STAT2 trial of high dose simvastatin in secondary PMS (SPMS) ^18^. MS-STAT2 was a multi-centre phase 3 trial in which 964 participants with SPMS were randomised 1:1 between simvastatin (80mg daily) or placebo across 31 sites in the United Kingdom (UK). Key eligibility criteria were age 25 to 65 years (inclusive), EDSS between 4.0-6.5 (inclusive) and a diagnosis of SPMS with steady progression within the preceding 2 years. 315 participants at a single trial site (University College London Hospital) were additionally enrolled into a sub-study measuring whole brain atrophy.Participants had 6 monthly visits for up to 4.5 years for the collection of CROMS, including the EDSS, T25FW and 9HPT. For participants in the sub-study WBA was measured from MRI at baseline and month 12, 24 and 36 of follow-up.

The MS-STAT2 trial found no evidence of benefit for simvastatin compared to placebo on the primary outcome of EDSS CDP (hazard ratio, simvastatin compared to placebo: 1.13 [95% confidence interval (CI) 0.91 to 1.39], p=0.26), nor on the secondary endpoint EDSS-Plus CDP ^19^. In the MRI sub-study no effect was seen on whole brain atrophy rate, with percent brain volume change per year of -0.72% in the placebo group, and -0.73% in the simvastatin group (adjusted mean difference, simvastatin compared to placebo: -0.02% per year (95% CI -0.17 to 0.14), p=0.845) ^13^.

We now describe our modelling approach in two stages: first the model for the clinically reported outcome methods, then the model for brain atrophy rates, and finally the joint correlation structure.

### 3.2 Multivariate mixed model for repeated measures outcomes

To model *q* = 1, …, *Q* continuous outcome measures with repeated measures over time (such as EDSS, 25FW and 9HPT in our OCTOPUS example), we propose using the following multivariate model with inclusion of random slopes and intercepts for each outcome:

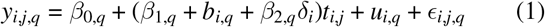

where

*y*_*i,j,q*_ is the measure for participant *i* = 1, …, *n* at visit *j* = 1, …, *J* for outcome *q* = 1, …, *Q*;

*δ*_*i*_ is the treatment group assignment; 1=active, 0=control;

*β*_0,*q*_ is the intercept for outcome *q*;

*β*_1,*q*_ is the shared mean slope for control participants for outcome *q*;

*β*_2,*q*_ is the difference in slope for those on active treatment versus control i.e. the treatment effect for outcome *q*;

*t*_*i,j,q*_ is the time value of the measure for participant *i* at visit *j* for outcome *q*;

*b*_*i,q*_ is the random slope for participant *i* for outcome *q*;

*u*_*i,q*_ is the random intercept for participant *i* for outcome *q*;

*ϵ*_*i,j,q*_ is the residual error for participant *i* at visit *j* for outcome *q*, and

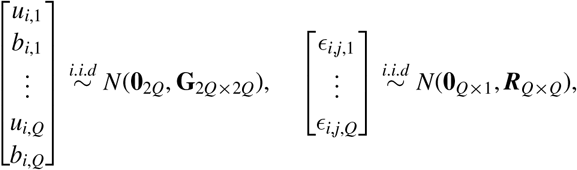

where **G, R**, are unstructured covariance matrix for the random effects and residuals respectively.

#### 3.2.1 Model for directly measured change outcomes

We now describe the model for the directly measured change in brain volume between pairs of visits; above we had *Q* continuous outcomes, so this is the (*Q* + 1)^*th*^ outcome measure. Let *y*_*i,j,k*,(*Q*+1)_ be the *directly measured* % difference in brain volume for participant *i* between visits *j* and *k*. We term this *directly measured* because each difference is separately determined by aligning the two brain scans. In consequence, for example, for times 0 < *j* < *k*, the difference in brain volume for patient *i* between times 0 and *k* will not exactly equal the difference from time 0 to time *j* plus the difference from time *j* to time *k*. In other words, *y*_*i*,0,*k*,(*Q*+1)_≠*y*_*i*,0,*j*,(*Q*+1)_ + *y*_*i,j,k*,(*Q*+1)_.

The formulation for a mixed model for this type of directly measured repeated data is described by Frost et al. ^20^. Its derivation from the underlying direct measurements means that, in addition to a random slope, the model includes visit specific random effects to allow for the expected correlation between directly measured changes that share either the start or end MRI scan, as well as independent difference specific residuals:

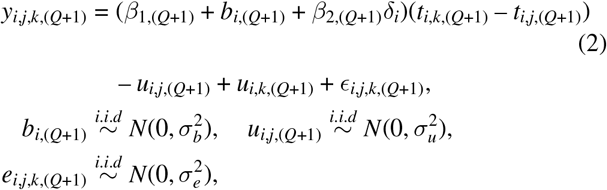

Here

*y*_*i,j,k*,(*Q*+1)_ is the direct difference measure for participant *i* (*i* = 1, …, *n*) between the *j*^*th*^ and *k*^*th*^ (1 ≤ *j* < *k ∈* 1, …, *J*) visit for outcome (*Q* + 1)

*δ*_*i*_ is the treatment group assignment; 1=active, 0=control

*β*_1,(*Q*+1)_ is the shared mean slope for control participants on outcome *q*

*β*_2,(*Q*+1)_ is the difference in slope for those on active treatment versus control i.e. the treatment effect for outcome *q*

*t*_*i,k*,(*Q*+1)_ – *t*_*i,j*,(*Q*+1)_ is the time difference between the pair of visits *k* and *j* for participant *i* forming the direct difference measure for outcome *q*

*b*_*i*,(*Q*+1)_ is the random slope for participant *i* for outcome *q*

*u*_*i,j*,(*Q*+1)_, *u*_*i,k*,(*Q*+1)_ are random subject-specific deviations at visits *j* and *k* for brain volume difference outcome (*Q* + 1)

*ϵ*_*i,j,k*,(*Q*+1)_ is the residual error for participant *i* on difference measure between visits *j* and *k* for brain volume difference outcome (*Q* + 1).

#### 3.2.2 Multivariate model for repeated measures and directly measured change outcomes

We link sub-models (1) and (2) by appending the random effects, *u*_*i,j*,(*Q*+1)_ from (2) to the vector of random effects from (1), so that

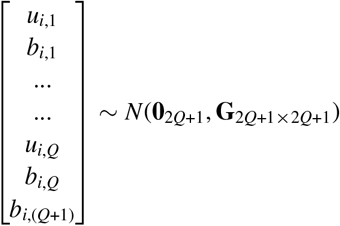

As before, **G** is unstructured, allowing for flexible modelling of the correlation between the the random slope for the directly measured change and the repeated measures of other outcomes.

The other random terms from (2) are independent from all other random terms from either sub-model and are distributed:

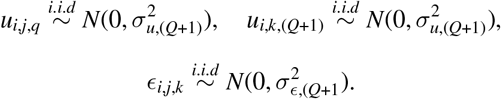

Note that V[*u*_*i,j*,(*Q*+1)_] = V[*u*_*i,k*,(*Q*+1)_].

#### 3.2.3 Hypothesis test for a treatment effect

From the multivariate mixed effects model of 3.2.2 we have a variance-covariance matrix of fixed parameters including a treatment effect (difference in rate of change) for each outcome.

To enable us to combine these treatment effects into a single measure we propose scaling the outcomes according to their respective random slope standard deviation. The model is fitted initially to obtain the random slope standard deviation and then fitted again following outcome rescaling. The effect is to provide standardisation according to the rate of change, so that unit treatment effects on the outcomes are broadly commensurate.

The proposed treatment effect is therefore a linear combination of the *β*_2*q*_ specified in the submodels(1) and (2) for the *q* = 1, …, (*Q* + 1) outcomes with weights {*c*_1_, *c*_2_, ..*c*_(*Q*+1)_}

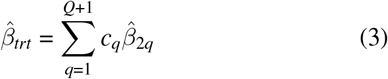

with its variance calculated as:

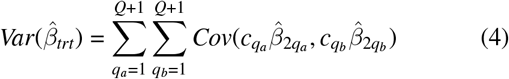

Different weighting schemes, such as inverse-variance weighting (IVW), could be utilised to combine the *Q* outcome effect estimates. However, as we have standardised the outcomes to a common scale of rate of change, an equal weighting approach can also be used. This restricts all weights to *c*_*q*_ = 1.

A hypothesis test can then be conducted against the null that 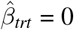 with the test statistic:

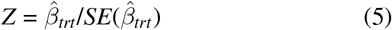

#### 3.2.4 Fitting the multivariate linear mixed model

The multivariate linear mixed model that simultaneously estimates sub-model equations (1) and (2) can be fit using restricted maximum likelihood using statistical packages for repeated measurements such as MLwiN ^21^.

There is still the possibility that the model does not converge, which may be a particular concern given potential data sparseness at the trial midpoint, and the many random (co-)variances to be estimated. In this situation we propose fitting the *Q* outcome sub-models separately. The variance-covariance matrix of the treatment effects may is then estimated using bootstrap-derived sampling distribution that respects the correlated nature of the data by sampling at the participant level within treatment arms.

### 3.3 Calculating power for a MAMS design

#### 3.3.1 Power for the multivariate mixed model

The general approach we use for calculating sample size or power for this model is based on methods described in Frost et al ^22^. First, we stack the outcomes for individuals to form an outcome vector **Y**, form the corresponding fixed effects design matrix **X**, random effects design matrix, **Z** and stack the level two random effects and residual vectors to write the model in the following form:

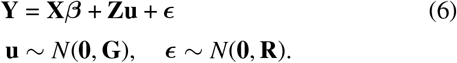

To perform the power calculation, following ^22^, we first derive the standard error of the treatment effect for a ‘2-patient’ trial (one active, one control); we then scale this by 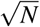 for a trial with *N* patients in each of two arms. To do this, we define the design matrices for the fixed effects **X** and random effects **Z** for the ‘2-person’ trial, with a given visit pattern and a single person in each treatment group.

The marginal form of this model is:

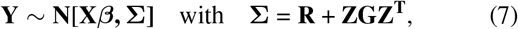

so that **Σ** is the marginal variance–covariance matrix for the ‘2-person’ repeated measures data **Y**.

For a given covariance matrix for the random effects **G** and for the residual error **R**, defined by the joint model for CROMS and brain atrophy set out above, the marginal variance–covariance matrix for the repeated measures of the outcome over time **Σ** can be calculated.

The variance-covariance matrix of the fixed effects, including all *Q* treatment effects, for the notional trial with one randomised pair of subjects, one in each arm (n=2), is then:

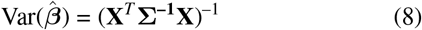

Hence Var(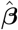) for a trial of size 2*N* (*N* in each of two arms) is

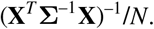

Next, we account for (assumed not intermittent) dropout and different visit patterns at interim analysis, as will happen in a MAMS trial due to staggered recruitment, by a pattern mixture approach ^23^. Suppose the focus is on the variance of the parameter estimates at a fixed time after ther trial opens. At this point, there will be cohorts of participants who have completed differing numbers of follow-up visits. We represent this by dividing the data into *m* cohorts based on the visit *m* = 1, .., *M* that the participant *i* has reached with probability *p*_*m*_, with *p*_1_ + .. + *p*_*M*_ = 1. In this setting, the overall estimate of 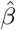 is the inverse-variance average of the estimates for each of the *m* cohorts:

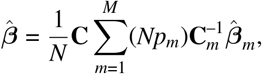

where 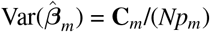 and we define

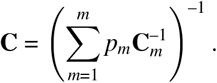

It follows:

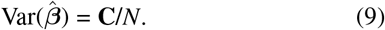

Finally, we may calculate the treatment effect and its variance for any linear combination of the elements of 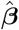; in particular, any (scalar) combination of the (*Q* + 1) treatment slope effects of interest, using equations (3), (4). Denote the variance of this estimate by *σ*^2^/*N*.

A sample size or power calculation can then be performed using standard approaches, for almost any linear mixed model, by this method. The required sample size per arm for power of 1 – *β* with significance level *α* (technically one-sided *α*/2) and difference ∆ may be computed as:

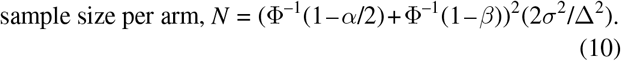

Alternatively, the power is

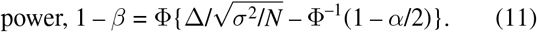

### 3.4 Calculating power for OCTOPUS using data from MS-STAT2

We demonstrate the method by applying it to the calculation of power for the new proposed OCTOPUS interim analysis.

#### 3.4.1 Analysis of data from MS-STAT2

We fitted the multivariate mixed model of 1 and 2 to the control arm only from the MS-STAT2 trial data for EDSS, T25FW and 9HPT and PBVC. Data were restricted to those visits up to the first 36 months and covariate adjustment was made for age, gender and baseline EDSS category. The 4 outcomes were standardised by dividing by each outcome by its respective random slope standard deviation, as previously described, and the model refitted to the standardised data. This model provided the estimates of the covariance matrix for the random effects **G** and for the residual error **R**

#### 3.4.2 OCTOPUS power calculation

The **G** and **R** matrices were then combined with appropriate **X** and **Z** design matrices for each visit-cohort *m* (as defined in 3.3.1) according to 7 to obtain the variance of the treatment effect under that visit pattern, as described in the previous section. The cohort weights *p*_*m*_ reflect the expected follow-up pattern in OCTOPUS at the time of the interim analysis, which was based on observed, and future expected, recruitment rates. The weights also reflect an assumed dropout pattern, based on a Weibull function with shape parameter=0.9 and a rate parameter back-calculated so that the proportion still in the trial at 3 years is 90%. For this joint model the design/variance matrices are not straightforward, and Supplement 1 provides a datasheet entry version of **X, R, Z, G** and Var(***β***_***m***_) for each cohort and the respective cohort weights used, as well as the estimated overall Var(***β***). As MRI is only being conducted for the first 125 participants in each arm, based on recruitment patterns it is anticipated approximately 215 participants will only have data on the clinically reported outcomes and hence for 3 cohorts the design matrices will reflect only sub-model 1.

The original power calculation for OCTOPUS Analysis Stage 1 (AS1) utilised a relaxed one-sided alpha of 0.35. We are prepared to increase this further to 40% or 45% in order to maintain a AS1 power of 0.9 and maximise the chance of detecting any worthwhile signal of treatment benefit. Plausible treatment effects used for power calculations consist of 10% and 15% reduction in the rate of brain atrophy (the latter equivalent to about 0.1%/year before standardisation) and 20%, 30% and 40% reduction in the rate of change for all CROMS, all relative to the respective control slopes seen in MS-STAT2. We also compared IVW with an equal-weighting approach, and used 0/1 (i.e. exclude/include) weightings to assess the individual performance of each of the 4 outcomes, and consideration of CROMS alone without atrophy.

These effect sizes we have assumed for OCTOPUS are deemed reasonable but are, naturally, unknowable prior to analysis. We therefore also compare if we instead use the relevant treatment effect sizes seen in the EXPAND ^17^ and ORATORIO ^14^ studies in the power calculation.

## 4 RESULTS

### 4.1 Illustration of potential power gains

Analysis of the control arm data from MS-STAT2 led to estimates of progression for each of the 4 outcomes based on the respective fixed slope effects, standardised relative the random slope variability. Only EDSS had a positive slope (0.42, standard error 0.058), whereas T25FW (-0.78, 0.069) and 9HPT (-0.34, 0.068) had negative slopes given they are speeds (which decline with disease progression) not times to complete a task. For whole brain atrophy decline is also measured as a decrease over time (-1.36, 0.11). These results suggest differential scope for possible improvement in absolute terms, and so support the proposal to specify effects in relative terms.

Table 2 provides power estimates based on the variance components derived from the main data source MS-STAT2, which are provided in Supplement 1. The calculations are given for each outcome measure in turn, for the 3 CROMS jointly, for all 4 outcomes jointly (both with equal-weighting) and for all 4 outcomes jointly with IVW. In each case the effect size and the alpha criterion is varied as described in Section 3.4.

**TABLE 2.** Power for differing outcomes, effect sizes and alpha criteria.

| Alpha | ES atrophy | ES CROMS | Atrophy | EDSS | T25FW | 9HPT | CROMS | C+A | C+A IVW |
| --- | --- | --- | --- | --- | --- | --- | --- | --- | --- |
| 0.35 | 10% | 20% | 0.633 | 0.552 | 0.646 | 0.477 | 0.645 | 0.707 | 0.707 |
|  | 10% | 30% | 0.633 | 0.652 | 0.775 | 0.543 | 0.774 | 0.807 | 0.806 |
|  | 10% | 40% | 0.633 | 0.742 | 0.872 | 0.607 | 0.871 | 0.883 | 0.882 |
|  | 15% | 20% | 0.758 | 0.552 | 0.646 | 0.477 | 0.645 | 0.753 | 0.755 |
|  | 15% | 30% | 0.758 | 0.652 | 0.775 | 0.543 | 0.774 | 0.843 | 0.844 |
|  | 15% | 40% | 0.758 | 0.742 | 0.872 | 0.607 | 0.871 | 0.908 | 0.908 |
| 0.4 | 10% | 20% | 0.681 | 0.604 | 0.694 | 0.530 | 0.693 | 0.750 | 0.751 |
|  | 10% | 30% | 0.681 | 0.699 | 0.813 | 0.594 | 0.812 | 0.841 | 0.840 |
|  | 10% | 40% | 0.681 | 0.783 | 0.898 | 0.656 | 0.897 | 0.907 | 0.906 |
|  | 15% | 20% | 0.797 | 0.604 | 0.694 | 0.530 | 0.693 | 0.793 | 0.795 |
|  | 15% | 30% | 0.797 | 0.699 | 0.813 | 0.594 | 0.812 | 0.873 | 0.873 |
|  | 15% | 40% | 0.797 | 0.783 | 0.898 | 0.656 | 0.897 | 0.928 | 0.928 |
| **0.45** | 10% | 20% | 0.725 | 0.652 | 0.737 | 0.580 | 0.737 | 0.789 | 0.789 |
|  | 10% | 30% | 0.725 | 0.742 | 0.845 | 0.643 | 0.844 | 0.870 | 0.869 |
|  | 10% | 40% | 0.725 | 0.818 | 0.919 | 0.702 | 0.918 | 0.926 | 0.925 |
|  | 15% | 20% | 0.831 | 0.652 | 0.737 | 0.580 | 0.737 | 0.828 | 0.829 |
|  | **15%** | **30%** | **0.831** | **0.742** | **0.845** | **0.643** | **0.844** | **0.897** | **0.898** |
|  | 15% | 40% | 0.831 | 0.818 | 0.919 | 0.702 | 0.918 | 0.944 | 0.944 |

C+A: CROMS+Atrophy
C+A IVW: CROMS+Atrophy with inverse-variance weighting
bold entries indicate final selected calculation inputs and power outputs

For the effect sizes chosen, relative to their variances, atrophy performs comparably to the individual CROMS: not as well as the 25FWT but better than EDSS and the 9HPT, which is clearly the least powerful individual outcome as the outcome with smallest change over time, due in part to the shallowest control group slope. Accordingly, when considering CROMS together, the linearly combined outcome performs exactly the same as the 25FWT alone, where the addition of EDSS and 9HPT at least compensates for the downweighting of the most ‘informative’ measure but adds no more to the joint effect size.

Adding atrophy to the CROMS, however, does improve power by between 3-10% depending on alpha and effect size values considered. With a 15% reduction in atrophy slope and 30% reduction for each CROMS we are able to achieve a power of 90% when alpha is stretched to 45%, compared to an atrophy-only estimate of 83% — a 7% increase (values highlighted in bold in Table 2). These were considered plausible effect sizes and are used for the comparison in the next section comparing against observed effect sizes from EXPAND and ORATORIO.

Given the standardisation of outcomes according to rate of change of standard deviation, use of IVW made almost no difference to estimated power compared to the equal-weighting of atrophy and CROMS, and so is not considered further here.

### 4.2 Application to examples

#### 4.2.1 Using examples to estimate power in OCTOPUS

We use the EXPAND and ORATORIO effect size estimates as a basis of comparison for calculating power in OCTOPUS. Table 3 presents the main power calculations from 4.1 (in bold) using MS-STAT variance parameter inputs and the selected effect sizes (15% in atrophy, 30% for all 3 CROMS) against values that have either or both effect size and variance input taken from these alternative trial data.

**TABLE 3.**
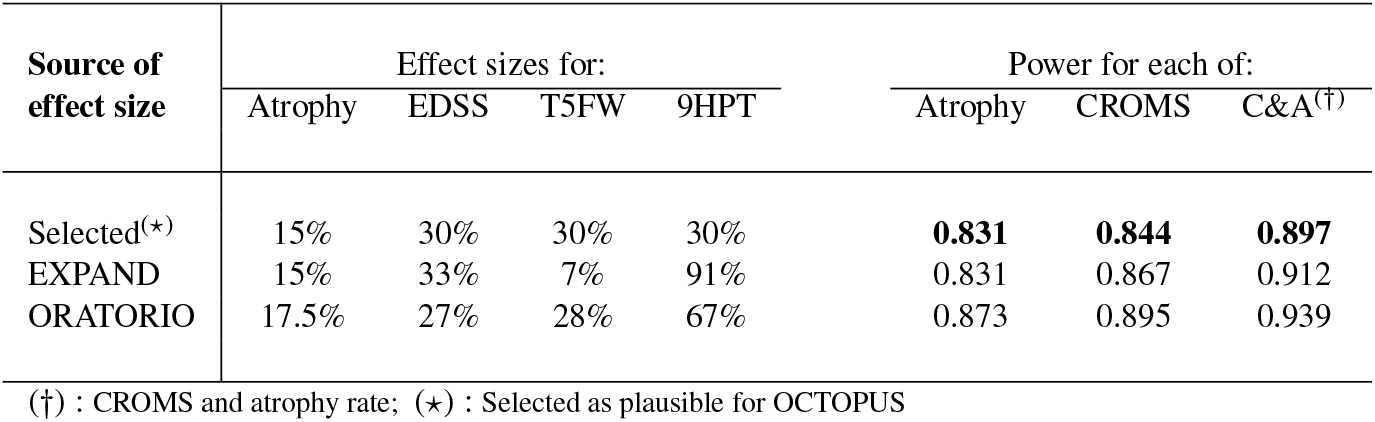
Power for differing sources of CROMS and atrophy treatment effect sizes (*α* = 0.45)

| Source of effect size | Effect sizes for: |  |  |  | Power for each of: |  |  |
| --- | --- | --- | --- | --- | --- | --- | --- |
|  | Atrophy | EDSS | T5FW | 9HPT | Atrophy | CROMS | C&A<sup>(†)</sup> |
| Selected<sup>(★)</sup> | 15% | 30% | 30% | 30% | **0.831** | **0.844** | **0.897** |
| EXPAND | 15% | 33% | 7% | 91% | 0.831 | 0.867 | 0.912 |
| ORATORIO | 17.5% | 27% | 28% | 67% | 0.873 | 0.895 | 0.939 |

(†) : CROMS and atrophy rate; (★) : Selected as plausible for OCTOPUS

The effect sizes seen in ORATORIO for atrophy, EDSS, 25FWT and 9HPT respectively are 17.5%, 27%, 28% and 67% — meaning they are similar to selected effect sizes except for 9HPT which is more than twice that value. For EXPAND the observed effect sizes are 15%, 33%, 7% and 91% respectively, so that for 25FWT it is considerably less than, and for 9HPT over three times as much as for the selected values. The overall impact to projected power in OCTOPUS AS1 with alpha held at 45% one-sided, is a small increase if the effects from EXPAND were seen, from 89.7% to 91.2%. If the effects in ORATORIO were seen, there is another moderate increase to 93.9%.

#### 4.2.2 Applying the new Analysis Stage 1 proposal to MS-STAT2

We applied the analysis model to the data from the MS-STAT2 trial. The results of the analysis showed simvastatin would have been dropped from the trial at the interim analysis as the combined test statistic for the four outcomes found there was no effect of the drug (-0.053 [95%CI -0.271, 0.165] p=0.634). Further, there was not a signal of the drug working on any of the four individual components. The effects on the individual components were: EDSS 0.020 [95%CI -0.244, 0.283] p=0.883; T25FW -0.048 [95%CI -0.336, 0.241] p=0.745; 9HPT -0.043 [95%CI -0.449, 0.362] p=0.834; WBA -0.101 [95%CI -0.485, 0.283] p=0.605]

## 5 DISCUSSION

For the interim stage analysis of MAMS studies, we need the most sensitive measure of possible drug activity against the disease: this does not to need to be a formal surrogate for the primary outcome, and unlike the primary outcome, it does not need a direct clinical interpretation. Therefore, at interim analyses, it makes sense to look to combine information on the various outcome measures to inform the decision to stop, or continue, with an arm.

In particular, as discussed in the introduction, our work was motivated by the OCTOPUS study in progressive MS, and in particular emerging concerns about the sensitivity of whole brain atrophy (WBA) to drugs that ultimately showed clinical efficacy.

The methodology described here provides a practical strategy for combining information on MRI WBA and CROMS, with the goal of increasing power for the OCTOPUS trial at Analysis Stage 1 and mitigating against the risk of unknown treatment effects for different outcomes. Although, ultimately, in this example the power gains have proven relatively modest, we believe the approach is an effective and generalisable one that may be adopted by any MAMS trial seeking to increase the probability of detecting a signal for candidate drug activity for an early stage analysis.

Our example based on OCTOPUS was particularly complex, as it not only involved the modelling of 4 joint outcomes but also the particular combination of three repeated clinical measure outcomes plus an imaging-based ‘directly measured’ outcome, resulting in a complicated analysis model and accompanying power calculation. However, regarding the overarching concept of combining multiple outcomes to calculate a test statistic and make stop/go decisions, the use of standard repeated continuous outcomes would be more likely. For this model3.2 the sample-size or power calculation can be performed with our user-written Stata command ‘mvmixedpower’, which is part of the more general ‘mixedpower’ package available at the SSC archive ^24^. The program allows the user to adjust for incomplete follow-up due to both staggered recruitment and dropout simultaneously, and has an option to automatically ‘read in’ the variance parameters from a suitable mixed model in memory into the calculation, which greatly saves on time and the chance of making a mistake versus entering a large covariance matrix manually. Also included in the package is ‘dmmixedpower’ which will similarly perform sample size and power calculations for a direct measures model 3.2.1.

Combining outcomes in this manner could limit the clinical interpretability of the treatment effect, which is why we suggest this as a strategy better suited for an interim analysis, which has less requirement for a meaningful effect size and greater emphasis on detecting a signal. When combined with a generous alpha criterion, potentially effective treatments are less likely to be dropped unnecessarily, whilst still providing the option for ineffective ones to be discontinued. A major MAMS trial for Parkinson’s, EJS ACT-PD ^25^, is now underway where the first 2 stages test a IVW combination of the 3 treatment effects estimated from a multivariate mixed model based on parts I, II and III of the MDS-UPDRS, which is the gold-standard clinical tool used to assess the severity and progression of Parkinson’s disease. For the final 2 stages testing for efficacy, an equal-weighted version of just part I and II will be used which has been declared a clinically valid summation by the scale authors ^26^.

Assuming our selected effect size inputs, including 30% effectiveness for CROMS and 15% for atrophy, the gain in power from adding the CROMS to atrophy was 7%, where maintaining 90% power at AS1 was achieved if alpha was raised to 45%. This was a considerable amount of work for an apparently moderate increase in power. However, there is always considerable uncertainty when stating likely effect sizes in trial design, and an important facet of the multivariate method is that it can mitigate against overestimated treatment effects on some outcomes by incorporating effects where targets were met or exceeded - essentially spreading the risk across outcomes. Naturally, each outcome will reflect different aspects of disease progression, and so one may expect the treatment effects to vary even though they may be assumed equal for the purpose of sample size calculation. Here, we assume 30% for all 3 CROMS, when the results of EXPAND and ORATORIO show variation in relative effect size by their respective active treatment arms. Interestingly, of these CROMS, where the 9HPT contributes the least for a given % effectiveness, the observed relative effect was twofold (ORATORIO) or threefold (EXPAND) of our assumed level; whereas the most seemingly informative CROMS (T25FW) had a particularly low relative effect in EXPAND. Hence, in Table 3 the multivariate approach helped to balance these discrepancies in the linear combination of effects leading to a consistent degree of improvement over atrophy alone.

In our example one would expect the CROMS to be more correlated with each other than with atrophy. This is borne out from the treatment effect covariance matrix of 8 (Supplement 1), where intra-CROMS correlations (all between 0.17 and 0.32 in magnitude) are all larger than inter-CROMS-atrophy correlations. Larger correlations mean there is less benefit to adding an outcome as they provide less distinct information. Therefore, it would not be a beneficial strategy to include many joint outcomes without consideration of their individual statistical as well as clinical merit. Whilst adding additional outcomes does not make the power calculation any less feasible (other than certain time-limiting issues), in terms of analysis, the model may become increasingly harder to fit, both in terms of machine runtime and in the likelihood of failure to converge. In such situations one may also adopt the approach described in 3.2.4 where each outcome is individually modelled and the necessary covariances obtained from the bootstrap sampling.

Our example also demonstrates that any set of continuous outcomes may be combined even if they do not appear initially compatible. By either standardising in some manner or using inverse-variance or inverse-covariance weighting of the treatment effects one may make them broadly commensurate. Prior summation of the outcomes into a single outcome is a more straightforward approach but still requires decisions on effective weighting, and leads to exclusion of information by casewise deletion, unless each outcome is exactly equivalently non-missing. For OCTOPUS, amongst the 3 selected CROMS, EDSS had the least amount of missing data, whereas by design atrophy had far less measurement visits and the its outcomes do even correspond with those of the CROMS. Interestingly, power for an individual outcome where some data is missing can be higher in the multivariate setting (but using e.g. 1,0,0,0 weighting) than when modelled univariately, as additional information may be exploited through the covariances. Finally, by using a multivariate model instead of outcome-summation, the simultaneous provision of outcome-specific treatment effects along with their correlations can also provide helpful insight in an IDMC setting.

## 6 CONCLUSION

MAMS studies need sensitive signals of early activity of the drug (or intervention) against the disease, in order to allow discontinuation of arms unlikely to be effective, while retaining high power to continue if there is a sign of activity. In MAMS trials for progressive MS, evidence suggests that relying on a single measure (whether it is brain atrophy or a particular clinically reported outcome measure (CROM)) is sub-optimal. To address this, we have described a modelling approach which allows us to combine information from brain atrophy and CROMS to define a test of disease activity which has increased power to detect drug activity against the disease in the early phase of follow-up. We have demonstrated the utility of this approach, including by applying it retrospectively to the recently completed STAT2 trial in PMS, where the method would have correctly recommended stopping. We believe this approach is now ready for wider use, and will be applying it to the interim Analysis Stage 1 of the ongoing OCTOPUS study.

## Data Availability

All data requests should be submitted to JC for consideration in the first
instance. Access to available fully anonymised data of MS-STAT2 might be granted 12 months after publication, after review by Jeremy Chataway and the sponsor (University College London). Requesters will be asked to complete an application form detailing specific requirements, rationale, and proposed use. A data-sharing agreement will need to be signed. Requested data will be made available, along with supporting documentation (eg, data dictionary) on a secure server.
Participants of MS-STAT2 have given consent for further research/collaborations with the data from the trial

## Abbreviations

MAMS: multi-arm multi-stage.

## CODE

Code is available at request from the authors and was crosschecked by independent coding from MB and JN

## AUTHOR CONTRIBUTIONS

MB, JN and RB contributed to the analysis and coding. All authors contributed to the manuscript.

## ACKNOWLEDGMENTS

Matthew Burnell, Rachel Burton, Sean Apap Mangion and Jennifer Nicholas are supported by a grant from the UK MS Society (grant reference number 135). James Carpenter is supported by MRC Unit grant MC_UU_00004/07 and the MRC Centre of Research Excellence in Clinical Trial Innovation (CCTI) in partnership with the NIHR, grant no. UKRI 934 .

